# EFFECT OF PNEUMOCOCCAL CONJUGATE VACCINE ON THE LENGTH OF HOSPITALIZATION IN CHILDREN UNDER FIVE WITH PNEUMONIA IN BURKINA FASO

**DOI:** 10.1101/2025.11.17.25340457

**Authors:** Mamadou Bountogo, Bintou Sanogo, Moïse Da, Haoua Tall, Caroline Martin, Windpanga Aristide Ouédraogo, Abdou-Salam Ouedraogo, Edouard Betsem, Nicolas Meda

## Abstract

**Background:** Pneumococcus is a leading cause of childhood morbidity and mortality worldwide. We assessed the impact of the 13-valent pneumococcal conjugate vaccine (PCV13) on the hospitalization length among children under five years hospitalized for pneumonia in Burkina Faso.

**Methods:** We conducted a secondary analysis of data collected from hospitalized pneumonia cases participating in a diagnostic validation study between December 2015 and January 2017. A Cox proportional hazards model was used to assess the association between vaccination status and time to discharge.

**Results:** A total of 280 children were included, of whom 187 (66.8%) had received three PCV13 doses. The mean hospitalization duration was 7.99 days (95% CI 7.17–8.81) for those fully vaccinated vs. 10.44 days (95% CI 9.07–11.80) among children with <3 doses. Full vaccination was associated with a significantly higher recovery rate (aHR: 1.45; 95% CI: 1.11–2.08, p = 0.01).

**Conclusion:** PCV13 significantly reduces hospitalization length due to pneumonia among children under five in Burkina Faso.

## INTRODUCTION

Streptococcus pneumoniae, a bacterium found in the human nasopharyngeal microbiota, is a pathogen that affects people of all ages, but is most prevalent in the very young and the very old (1). Pneumococcus is currently the leading cause of invasive bacterial infections in children aged three months to two years. The main disease caused by pneumococcus in children are pneumonia, bacteraemia, meningitis, otitis, and arthritis (2). Globally, in 2021, there were an estimated 7 million cases of pneumonia (3), including 502,000 deaths in children under 5 years of age (4). In Burkina Faso, pneumonia was the leading cause of medical consultations and the third leading cause of hospitalization in health services in 2022 (5). The average direct cost of hospitalization for pneumonia in paediatric patients ranges from US$115 to US$345 (6) (7).

In a 2012 summary note, the World Health Organization (WHO) recommended the introduction of pneumococcal conjugate vaccines into national immunization programs in countries around the world (7). From 2015 to 2021, the number of deaths from pneumonia in children under 5 around the world fell by 202,000 cases, a reduction of 28% (3,8). In sub-Saharan African countries, the declines were smaller, less than 12% for the number of pneumonia cases and 20% for mortality (3). In Burkina Faso, the 13-valent vaccine was introduced in October 2013(9).

According to the recommendations of the Global Immunization Monitoring and Surveillance Framework, countries introducing vaccines such as the pneumococcal conjugate vaccine (PCV) should evaluate the effectiveness of vaccination on the occurrence of diseases targeted by these vaccines (10). In Burkina Faso, several studies have been conducted on mortality, the incidence of pneumonia, and nasopharyngeal carriage of pneumococcus after the introduction of PCV (9,11,12). In this study, we propose an assessment of the impact of PCV-13 on the length of hospitalization in children hospitalized for pneumonia in Burkina Faso.

## MATERIALS AND METHODS

### Study site

This study was conducted in the three main hospitals in Bobo-Dioulasso where cases and their controls were recruited: Sourô Sanou University Hospital (CHUSS), Dô Medical Center with Surgical Unit (CMA), and Dafra CMA.

The CHUSS is the university hospital for the western part of Burkina Faso. This hospital has a threefold mission: care, training, and research. It is located in Bobo-Dioulasso city. The CHUSS was the main clinical site for the identification and inclusion of hospitalized patients. The CMA of Dô is the hospital that serves the Health and Social Promotion Centers (CSPS) in the Dô health district. This hospital has a large capacity, and the number of pneumonia cases hospitalized in 2014 was estimated at 800. The CMA of Dô is located about 4 km from the CHUSS. The CMA of Dafra is the referral hospital for the CSPSs in the Dafra, Lena, and Karangasso-Vigué health districts. The CMA of Dafra is located approximately 5 km from the CHUSS.

### Study period

The data collection took place from December 2015 to January 2017.

### Study design

This was a survival analysis of data collected as part of a study that aimed to determine the appropriate detection thresholds for a urinary pneumococcal antigen diagnostic test and its use in a case-control study to assess the impact of PCV-13 in children under 5 years of age in Burkina Faso.

### Participant enrolment

Children under five years of age who were hospitalized with pneumonia were eligible for inclusion. Recruitment was conducted daily between 7:00 a.m. and 5:00 p.m. at the three participating hospitals, following a physician-confirmed diagnosis of pneumonia. The study employed an exhaustive sampling strategy that included all hospitalized pneumonia cases among children under five years of age across the three sites.

Clinical personnel at each hospital received standardized training on study procedures, and a designated physician at every site supervised participant enrolment to ensure protocol adherence. Children admitted during night hours were screened and enrolled the following morning. Eligibility criteria were: (i) hospitalization of a child aged under 5 years; (ii) written informed consent obtained from a parent or legal guardian; and (iii) fulfilment of the World Health Organization (WHO) clinical case definition of pneumonia (https://iris.who.int/bitstream/handle/10665/43551/9789242546705_fre.pdf.

### Data Collection

Information on each child’s medical history, socio-demographic, and household characteristics were collected using a standardized questionnaire. Vaccination data were collected from the child’s vaccination card and children without a documented vaccination record were excluded from the analysis

### Variables

In this study, the primary outcome, exposure and potential confounding variables were clearly defined prior to analysis, in accordance with STROBE recommendations. Table 1 provides an overview of all variables included in the analysis, including their operational definitions, sources of data, and coding procedures used for statistical modeling. The outcome variable was time to hospital discharge, defined as the duration of hospitalization until discharge following clinical recovery. The main exposure was the child’s PCV-13 vaccination status. Additional covariates capturing demographic, clinical, environmental and socioeconomic characteristics were considered as potential confounders based on literature and clinical relevance.

**Table I:** variables and category.

| Variable | Type | Definition / Measurement | Coding (analysis) | Source | STROBE Note |
| --- | --- | --- | --- | --- | --- |
| Time to discharge (days) | Outcome | Duration from admission to discharge (days). | Continuous time; event indicator: 1 = discharged recovered; 0 = death / transfer / discharge against medical advice. | Hospital records (admission/discharge notes). | Clearly define outcomes; specify coding of events/censoring. |
| PCV-13 vaccination status | Exposure | Receipt of 3 doses of PCV-13 before admission. | 1 = 3 doses (fully vaccinated); 0 = <3 doses (incompletely vaccinated). | Child vaccination card / health record. | Clearly define exposures and ascertainment method. |
| Age group (months) | Covariate | Age at admission in completed months; categorized as in analysis. | ≤15; 16–30; 31–45; ≥46 (reference group specified in models). | Hospital records / caregiver report. | Report how categories were chosen. |
| Sex | Covariate | Biological sex of child. | Male / Female. | Hospital records. | Describe demographics. |
| Place of residence | Covariate | Usual residence of the household. | Urban / Rural. | Questionnaire. | Contextual characteristic. |
| MUAC | Covariate | Mid-upper arm circumference at admission. | $\geq 125$ mm (Ref) / $< 125$ mm. | Clinical assessment. | Potential confounder; report measurement method. |
| Number of rooms in household | Covariate | Total distinct rooms in the dwelling. | $> 2$ rooms (Ref) / $\leq 2$ rooms. | Questionnaire. | Socioeconomic proxy. |
| Kitchen located in sleeping area | Covariate | Whether the main cooking area is in the child's sleeping room. | Yes / No. | Questionnaire. | Indoor air exposure. |
| Use of fumigant substances | Covariate | Use of mosquito coils or similar in child's room. | Yes / No. | Questionnaire. | Indoor air exposure. |
| Cooking energy source | Covariate | Primary household fuel for cooking. | Wood/Charcoal (Ref) / Butane gas. | Questionnaire. | Environmental exposure. |
| Mother's education level | Covariate | Highest education attained by mother. | None; Primary; Secondary or higher. | Questionnaire. | Socioeconomic status indicator. |
| Father's education level | Covariate | Highest education attained by father. | None; Primary; Secondary or higher. | Questionnaire. | Socioeconomic status indicator. |
| Mother's occupation | Covariate | Main economic activity of mother. | Self-employed/Salaried; Housewife. | Questionnaire. | Socioeconomic status indicator. |
| Father's occupation | Covariate | Main economic activity of father. | Unemployed; Salaried; Self-employed. | Questionnaire. | Socioeconomic status indicator. |
| Adults sleeping in same room as child | Covariate | Number of adults sharing child's sleeping room. | >2 adults (Ref) / ≤2 adults. | Questionnaire. | Crowding indicator. |
| Referred from another facility | Covariate | Referral status at admission. | Yes / No. | Hospital records. | Severity proxy. |
| Convulsions | Covariate | Convulsions at or before admission. | Yes / No. | Clinical notes. | Severity proxy. |
| Prostration / lethargy | Covariate | Clinical prostration or lethargy at admission. | Yes / No. | Clinical notes. | Severity proxy. |

The primary event of interest was hospital discharge. Discharge following clinical recovery was coded as 1, whereas discharge for any other reason—death, transfer to a higher-level facility, or discharge against medical advice—was coded as 0. The outcome variable was defined as the time discharge (i.e., duration of hospitalization until the occurrence of the event of interest). The main explanatory variable was PCV-13 vaccination status. A child was considered *fully vaccinated* (coded as 1) if three doses of the PCV13 vaccine had been received, and *incompletely vaccinated* (coded as 0) if fewer than three doses (<3 doses) had been administered. All other covariates represented potential confounding factors.

### Quality control

Data collection was supervised by a clinical research assistant who verified the consistency of questionnaire data with each child’s hospital and vaccination records. Data were entered into *OpenClinica* and subjected to continuous quality checks. Inconsistencies triggered data queries, which were reviewed and corrected after verification against source documents.

### Data analysis

We used the Cox proportional hazards model to assess factors associated with time to hospital discharge. The model estimates the hazard function *h*(*t*|*x*) as the product of a baseline hazard function *h*_0_(*t*), which depends only on time, and a covariate function dependent on explanatory variables:

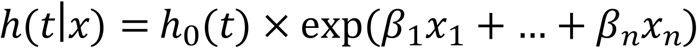

Model assumptions were verified by assessing log-linearity, checking proportional hazards graphically, and testing model fit using the likelihood-ratio test. All analyses were conducted using *Stata* version 14. Statistical significance was set at *p* < 0.05.

## RESULTS

Between December 2015 and January 2017, 280 children hospitalized with pneumonia across the three study hospitals were enrolled. Among these, 93 (35.2%) had received fewer than three doses of PCV-13, and 187 (66.8%) had received all three doses. Overall, children who had received three doses were older than those with fewer doses (p <0.001). Female children accounted for 34.4% of those with fewer than three doses and 35.3% of those fully vaccinated. Among children with fewer than three doses, 26.9% of mothers had at least primary school education, compared with 25.1% among those with three doses (Table II). Fathers had no formal education in 71% of cases among children with fewer than three doses, compared with 70.1% among those fully vaccinated. Prostration was observed in approximately 10% of children in both groups. More than 90% of patients were discharged after recovery in both groups.

**Table II:** Characteristics of children aged 0-59 months hospitalized for pneumonia from 2015 to 2017 according to vaccination status in three hospitals in the city of Bobo-Dioulasso, Burkina Faso.

|  | < 3doses |  | 3doses |  | P-value (3doses / <3doses) |
| --- | --- | --- | --- | --- | --- |
| Variable | N=93 | % | N=187 | % |  |
| Age group |  |  |  |  | 0.0001 |
| ≤ 15 months | 33 | 35.5 | 10 | 5.4 |  |
| 16–30 months | 20 | 21.5 | 58 | 31.0 |  |
| 31–45 months | 15 | 16.1 | 87 | 46.5 |  |
| ≥ 46 months | 25 | 26.9 | 32 | 17.1 |  |
| Sex |  |  |  |  | 0.88 |
| Male | 61 | 65.6 | 121 | 64.7 |  |
| Female | 32 | 34.4 | 66 | 35.3 |  |
| Place of residence |  |  |  |  | 0.70 |
| Rural | 43 | 46.2 | 82 | 43.9 |  |
| Variable | N=93 | % | N=187 | % |  |
| Urban | 50 | 53.8 | 105 | 56.2 |  |
| **MUAC** |  |  |  |  | **0.10** |
| ≥125mm | 70 | 75.3 | 156 | 83.4 |  |
| <125mm | 23 | 24.7 | 31 | 16.6 |  |
| **Number of rooms in the household** |  |  |  |  | **0.16** |
| > 2 rooms | 42 | 45.2 | 101 | 54.0 |  |
| ≤ 2 rooms | 51 | 54.8 | 86 | 46.0 |  |
| **Kitchen located in the child's sleeping area** |  |  |  |  | **0.42** |
| Yes | 3 | 3.2 | 10 | 5.4 |  |
| No | 90 | 96.8 | 177 | 94.7 |  |
| Variable | N=93 | % | N=187 | % |  |
| **Use of fumigant substances in the child's room** |  |  |  |  | **0.70** |
| No | 51 | 54.8 | 107 | 57.2 |  |
| Yes | 42 | 45.2 | 80 | 42.8 |  |
| **Cooking energy source** |  |  |  |  | **0.84** |
| Wood/Charcoal | 89 | 95.7 | 178 | 95.2 |  |
| Butane gas | 4 | 4.3 | 9 | 4.8 |  |
| **Mother's education level** |  |  |  |  | **0.88** |
| None | 68 | 73.1 | 140 | 74.9 |  |
| Primary | 15 | 16.1 | 26 | 13.9 |  |
| Secondary or higher | 10 | 10.8 | 21 | 11.2 |  |
| **Father's education level** |  |  |  |  | **0.98** |
| Variable | N=93 | % | N=187 | % |  |
| None | 66 | 71.0 | 131 | 70.1 |  |
| Primary | 16 | 17.2 | 34 | 18.2 |  |
| Secondary or higher | 11 | 11.8 | 22 | 11.7 |  |
| **Mother's occupation** |  |  |  |  | **0.45** |
| Self-employed / Salaried | 62 | 65.6 | 115 | 61.0 |  |
| Housewife | 33 | 34.4 | 74 | 39.1 |  |
| **Father's occupation** |  |  |  |  | **0.636** |
| Unemployed | 24 | 25.8 | 39 | 20.9 |  |
| Salaried | 60 | 64.5 | 130 | 69.5 |  |
| Self-employed | 9 | 9.7 | 18 | 9.6 |  |
| **Adults sleeping in the same room as the child** |  |  |  |  | **0.16** |
| Variable | N=93 | % | N=187 | % |  |
| > 2 adults | 91 | 97.9 | 176 | 94.1 |  |
| ≤ 2 adults | 2 | 2.2 | 11 | 5.9 |  |
| **Length of hospital stay (days)** | 93 | 10.44 (9.07–11.80) | 187 | 7.99 (7.17–8.81) | **0.005** |
| **Prostration or lethargy** |  |  |  |  | 0.87 |
| No | 83 | 89.3 | 168 | 89.8 |  |
| Yes | 10 | 10.8 | 19 | 10.2 |  |
| **Referred from another health facility** |  |  |  |  | **0.95** |
| No | 32 | 34.4 | 65 | 34.8 |  |
| Yes | 61 | 65.6 | 122 | 65.2 |  |
| **Convulsion** |  |  |  |  | **0.52** |
| No | 88 | 94.6 | 180 | 96.3 |  |
| Yes | 5 | 5.4 | 7 | 3.7 |  |
| Variable | N=93 | % | N=187 | % |  |
| **Outcome at discharge** |  |  |  |  | **0.88** |
| Recovered | 86 | 92.5 | 172 | 92.0 |  |
| Escaped, deceased, or transferred | 7 | 7.5 | 15 | 8.0 |  |

\* MUAC= Mid-upper arm circumference

The mean length of hospitalization was 7.99 days (95% CI 7.17–8.81) among children who had received three doses, compared with 10.44 days (95% CI 9.07–11.80) among those with fewer than three doses (Figure 1). More than half (over 50%) of children who received three doses were discharged after recovery by day 6 of hospitalization, whereas this proportion was below 25% among those with fewer than three doses (Figure 2). By day 12, fewer than 25% of fully vaccinated children remained hospitalized, whereas this threshold was reached only after 15 days among those with fewer than three doses.

**Figure 1:**
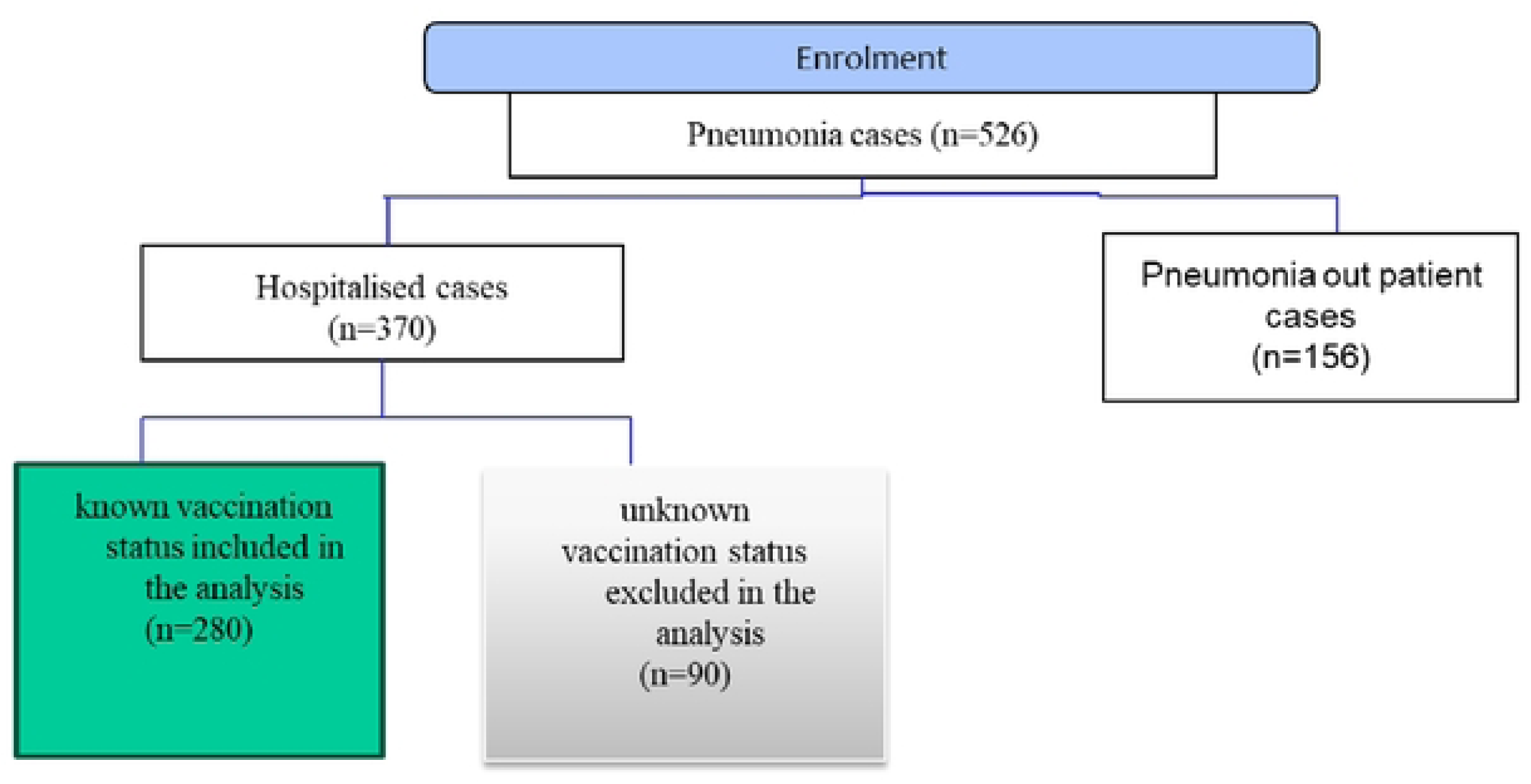
Enrolment flowchart of hospitalized pneumonia cases in th.is study.

**Figure 2:**
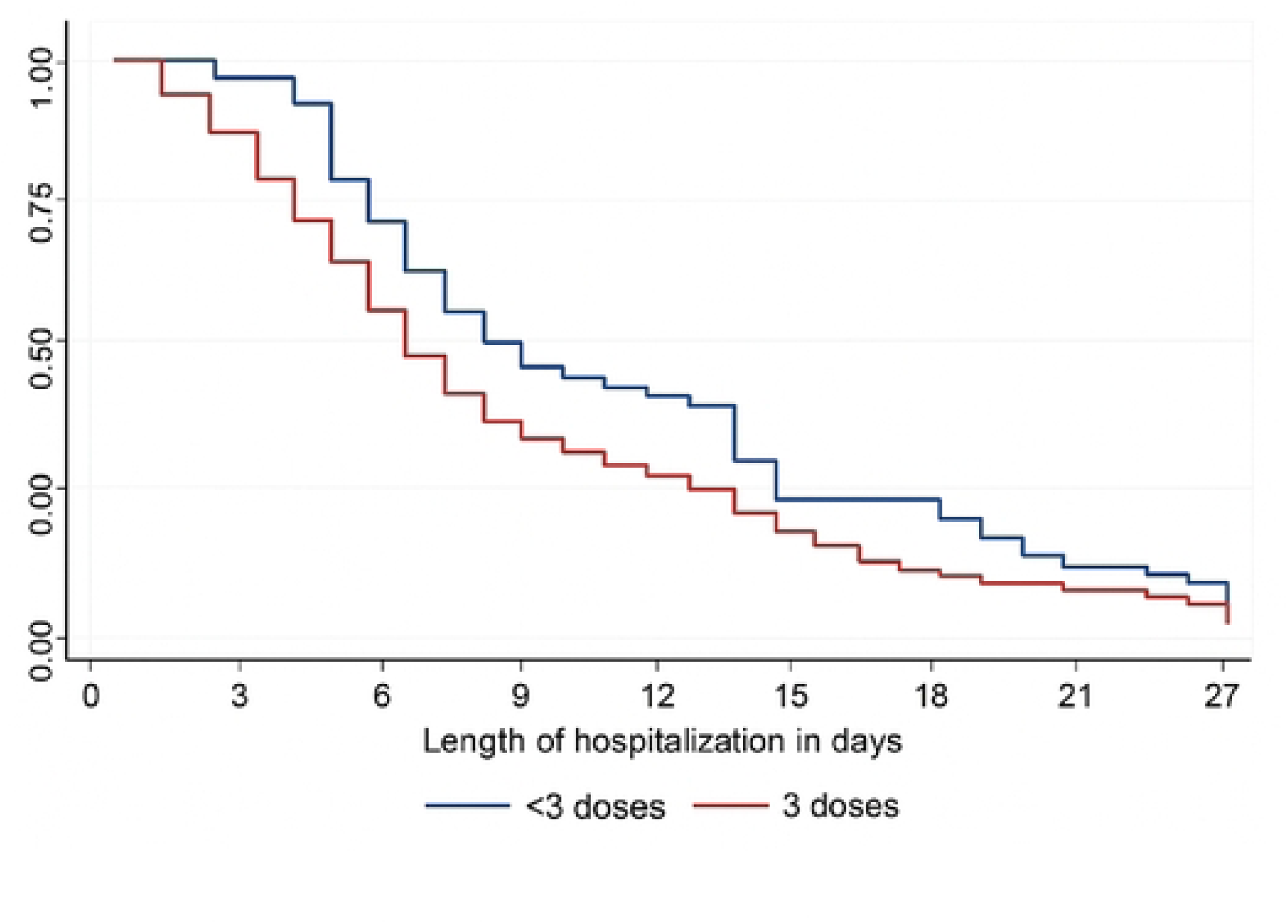
Kaplan Meier Survival curve of length of hospitalization by PCV-13stautt (<3dosesvs3dose) in children hospitalized for pneumoia, Burkina Faso

No clear difference in length of hospitalization was observed among children younger than 46 months, regardless of vaccination status (Figure 3). However, among those aged 46 months or older, the length of hospitalization was consistently shorter in the fully vaccinated group. By day 9 of hospitalization, more than 75% of children with three doses had been discharged after recovery, compared with fewer than 25% among those with fewer doses.

**Figure 3.**
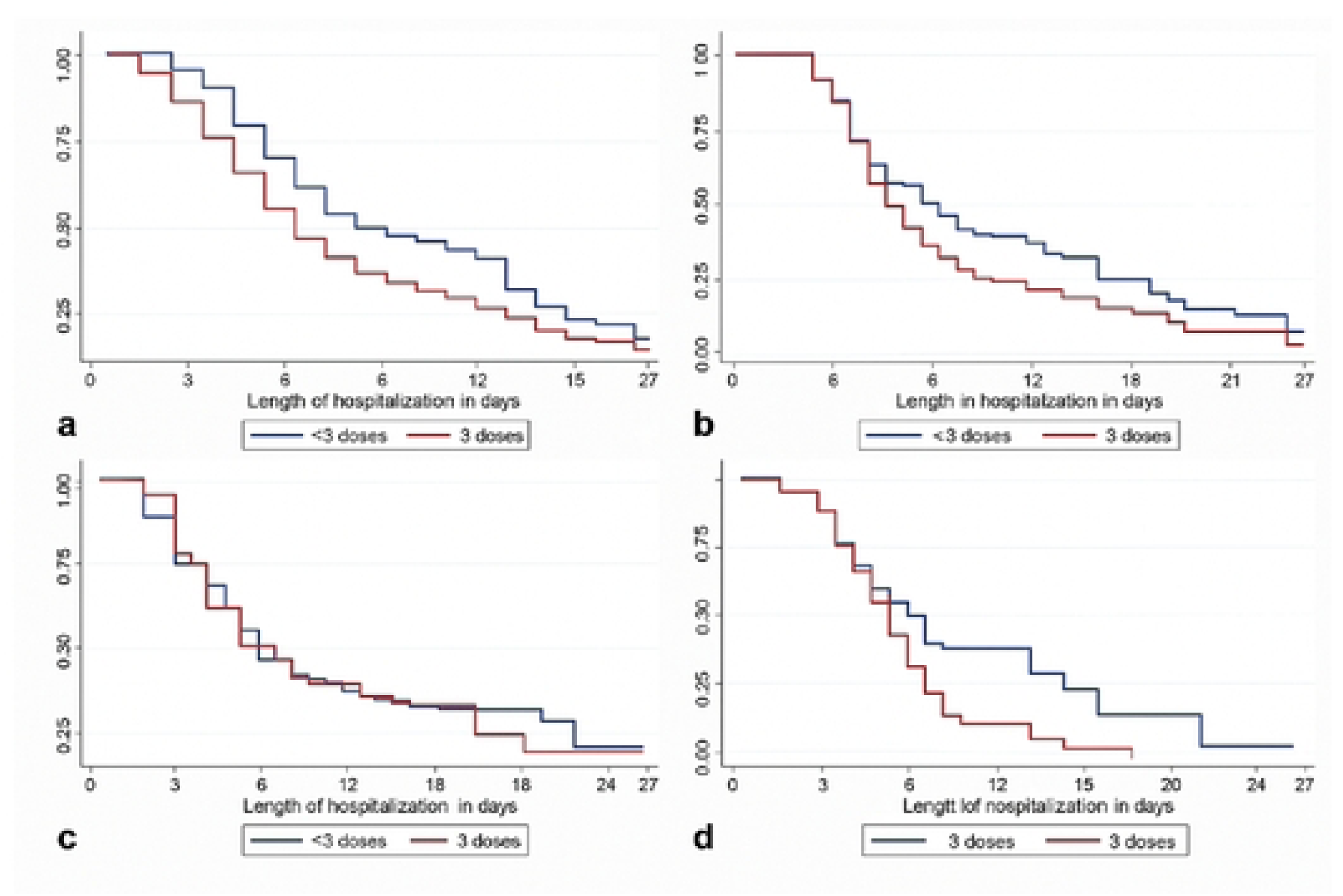
Estimated proportion of children hospitalised for pneumonia according to length of hospital stay, PCV-13 vaccination status, and age group in the three hospitals of Bobo-Dioulasso, Burkina Faso a) 0-15 months b) 16-30 months c) 31-45 months d) 46-59 months

The hazard ratio for released cured among children who had received three doses was 1.45 (95% CI 1.11–2.08; p = 0.01) (Table III). Thus, the instantaneous rate of recovery was 1.45 times higher among fully vaccinated children compared with those who had received fewer than three doses of PCV-13. Children with acute malnutrition had a 1.71-fold higher hazard compared with those with normal nutritional status (p = 0.002).

**Table III:**
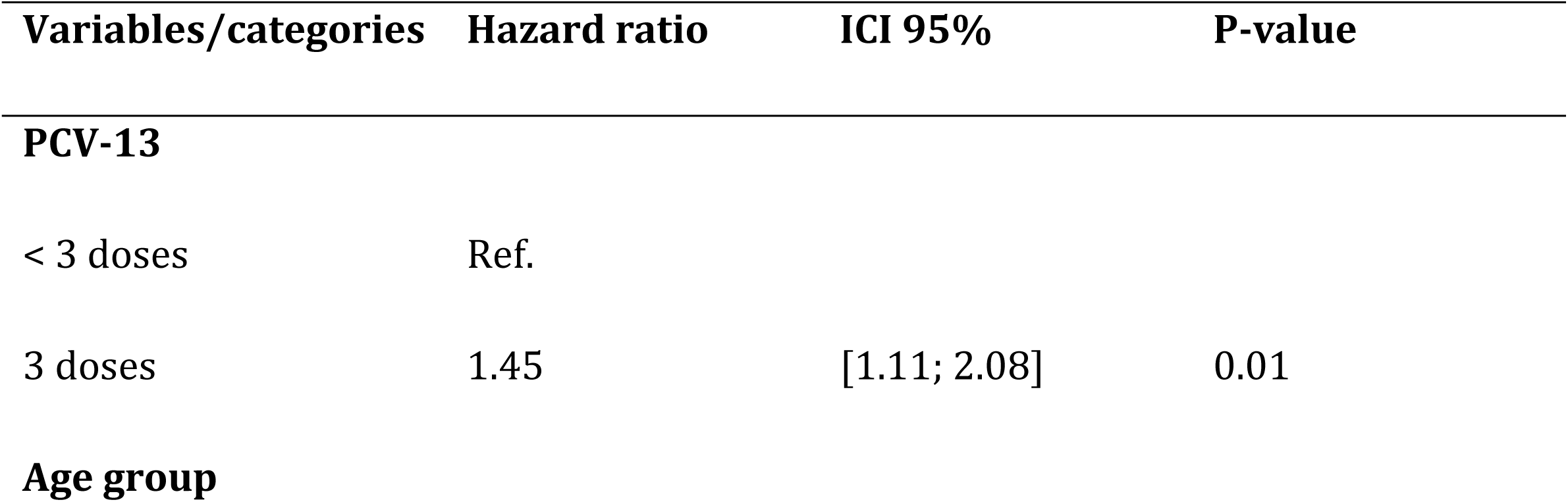
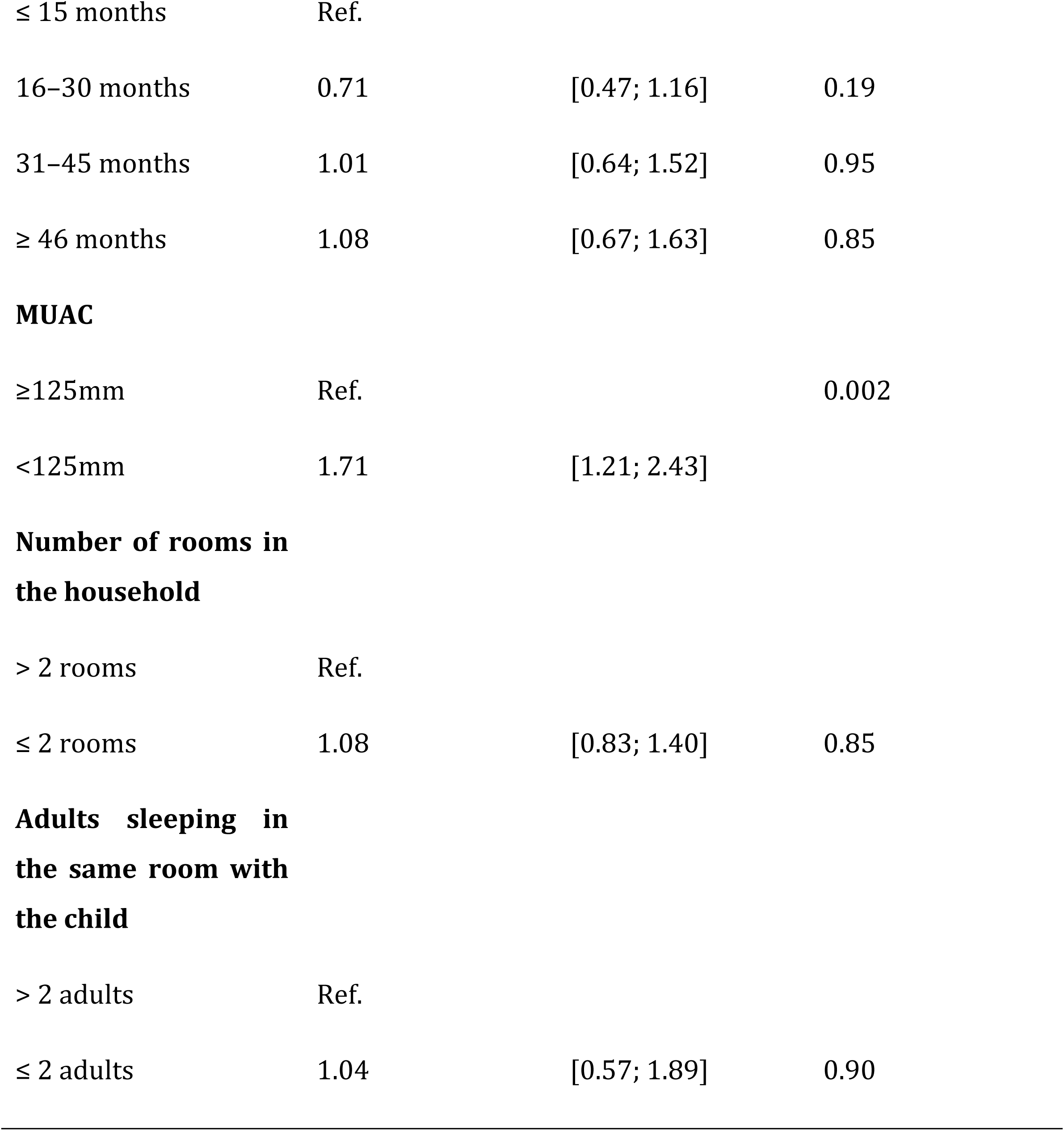
Cox models for the length of hospital stay of children aged 0-59 hospitalized for pneumonia in the three hospitals in the city of Bobo-Dioulasso, based on the doses of VCP-13 received and covariates.

| Variables/categories | Hazard ratio | ICI 95% | P-value |
| --- | --- | --- | --- |
| **PCV-13** |  |  |  |
| < 3 doses | Ref. |  |  |
| 3 doses | 1.45 | [1.11; 2.08] | 0.01 |
| **Age group** |  |  |  |
| ≤ 15 months | Ref. |  |  |
| 16–30 months | 0.71 | [0.47; 1.16] | 0.19 |
| 31–45 months | 1.01 | [0.64; 1.52] | 0.95 |
| ≥ 46 months | 1.08 | [0.67; 1.63] | 0.85 |
| **MUAC** |  |  |  |
| ≥125mm | Ref. |  | 0.002 |
| <125mm | 1.71 | [1.21; 2.43] |  |
| **Number of rooms in the household** |  |  |  |
| > 2 rooms | Ref. |  |  |
| ≤ 2 rooms | 1.08 | [0.83; 1.40] | 0.85 |
| **Adults sleeping in the same room with the child** |  |  |  |
| > 2 adults | Ref. |  |  |
| ≤ 2 adults | 1.04 | [0.57; 1.89] | 0.90 |

In linear regression analysis, children who received three doses of PCV-13 had a significantly shorter hospital stay compared with those who received fewer than three doses (β = –5.78, p = 0.002). Receiving only one or two doses was not significantly associated with a reduction in length of stay. (Table IV)

**Table IV.** Comparison of average hospital stays based on the number of VPC-13 received.

| **Length of hospital stay** | **slope** | **Standard deviation** | **P-value** |
| --- | --- | --- | --- |

| Dose |  |  |  |
| --- | --- | --- | --- |
| 0dose | Ref. |  |  |
| 1 dose | -5.168269 | 3.442892 | 0,134 |
| 2 doses | -3.770769 | 2.930922 | 0,199 |
| 3 doses | -5.784245 | 1.888022 | 0,002 |
| _intercept | 15.73077 | 1.670048 |  |

## DISCUSSION

In our study population, the mean length of hospital stays among children who received three doses of PCV-13 was 7,99. days (95%CI7,61-9.17). This duration is comparable to that reported by Kane et al. in Mali, who found an average of 9 days in a 2017 study on children with acute community-acquired pneumonia (13). Similarly, in Morocco Lahlimi et al. (2009–2011) observed an average hospital stay of 5.58±3.93 days for pneumonia in children under five years (14). These findings are consistent with ours. A study conducted in France (2014-2022), which compared data before and after the 2017 expansion of PCV-13 vaccination recommendations to at-risk individuals, reported an average of hospital stay of 9.8 days for invasive pneumococcal diseases (15). This difference is expected, as that study included all age groups and focused on invasive infections typically requiring prolonged hospitalisation.

In our cohort, children without malnutrition were 1,7 times more likely to be recovered any time during hospital stay than those with moderate malnutrition (p=0.002). Nutritional status is a well-established comorbidity that prolongs hospitalisation regardless of the underlying illness (16). In Niger, Kangaye et al. (2016) found that malnutrition increased the average length of hospital stay by approximately 45%, consistent with our observations. Malnutrition impairs immune function, making children more vulnerable to infections and slowing recovery (17).

Regarding vaccination status, children who received three doses of PCV-13 were 1.45 times more likely to be recovered than those than those who received fewer than three doses. Thus, receiving fewer than three doses reduced the likelihood of recovery—defined as survival without transfer or discharge against medical advice—by 45% during hospitalisation among children under five years.

This study was conducted two years after PCV-13 was introduced into Burkina Faso’s Expanded Programme on Immunization; therefore, its effect may have been partially attenuated by the indirect (herd) effects of conjugate vaccination (9,18). We compared the impact of full versus partial vaccination on hospital stay duration. Although this comparison could dilute the relative effect size, the association remained statistically and clinically significant.

In the primary survival analysis using the cox proportional hazards model, full vaccination was associated with a significantly higher rate of released cured (adjusted hazard ratio[aHR] 1.45).

## CONCLUSION

PCV-13 significantly reduced the length of hospitalization due to pneumonia among children under five years of age in Burkina Faso. This beneficial effect was strongest among older children close to five years of age. Continued high coverage of PCV-13 is warranted to sustain and strengthen this impact.

## Author Contributions

^¶^These authors contributed equally to this work.

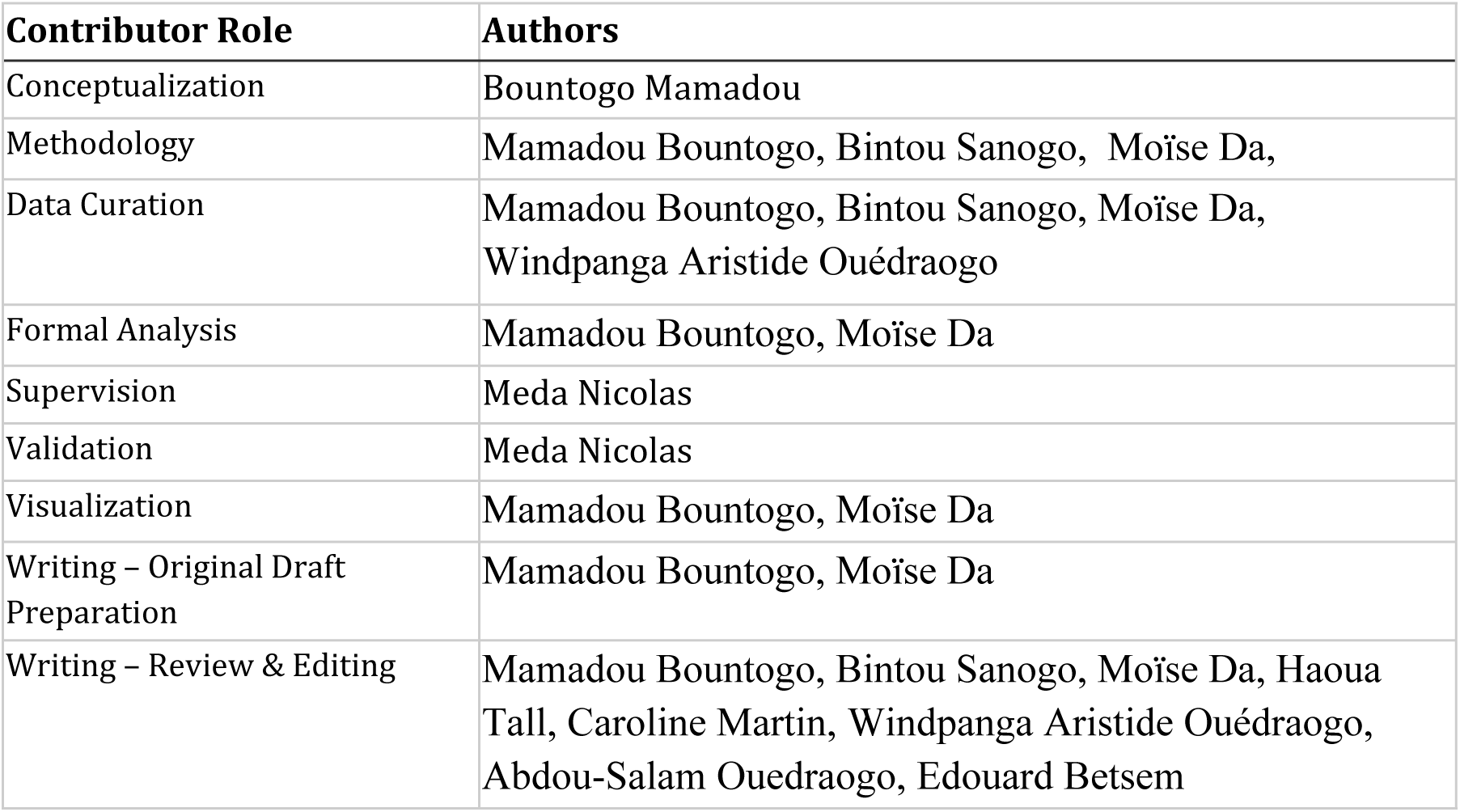

| Contributor Role | Authors |
| --- | --- |
| Conceptualization | Bountogo Mamadou |
| Methodology | Mamadou Bountogo, Bintou Sanogo, Moïse Da, |
| Data Curation | Mamadou Bountogo, Bintou Sanogo, Moïse Da,<br>Windpanga Aristide Ouédraogo |
| Formal Analysis | Mamadou Bountogo, Moïse Da |
| Supervision | Meda Nicolas |
| Validation | Meda Nicolas |
| Visualization | Mamadou Bountogo, Moïse Da |
| Writing – Original Draft Preparation | Mamadou Bountogo, Moïse Da |
| Writing – Review & Editing | Mamadou Bountogo, Bintou Sanogo, Moïse Da, Haoua Tall, Caroline Martin, Windpanga Aristide Ouédraogo, Abdou-Salam Ouedraogo, Edouard Betsem |

All authors have read and agreed to the published version of the manuscript.

## Ethics Statement

The study protocol was approved by the Burkina Faso Health Research Ethics Committee (Approval No. 2015-5-064, May 6, 2015). Written informed consent was obtained from parents/legal guardians.

## Data Availability

All relevant data are available and will be deposited in an open-access public repository prior to publication.

